# Representation learning for multi-modal spatially resolved transcriptomics data

**DOI:** 10.1101/2024.06.04.24308256

**Authors:** Kalin Nonchev, Sonali Andani, Joanna Ficek-Pascual, Marta Nowak, Bettina Sobottka, Tumor Profiler Consortium, Viktor H Koelzer, Gunnar Rätsch

## Abstract

Spatial transcriptomics enables in-depth molecular characterization of samples on a morphology and RNA level while preserving spatial location. Integrating the resulting multi-modal data is an unsolved problem, and developing new solutions in precision medicine depends on improved methodologies. Here, we introduce *AESTETIK*, a convolutional deep learning model that jointly integrates spatial, transcriptomics, and morphology information to learn accurate spot representations. *AESTETIK* yielded substantially improved cluster assignments on widely adopted technology platforms (e.g., 10x Genomics™, NanoString™) across multiple datasets. We achieved performance enhancement on structured tissues (e.g., brain) with a 21% increase in median ARI over previous state-of-the-art methods. Notably, *AESTETIK* also demonstrated superior performance on cancer tissues with heterogeneous cell populations, showing a two-fold increase in breast cancer, 79% in melanoma, and 21% in liver cancer. We expect that these advances will enable a multi-modal understanding of key biological processes.

## 1 Introduction

In multicellular organisms, cells are organized into tissues, groups of cells exhibiting common characteristics related to the biological function [1, 2]. Recent advances in spatial transcriptomics enable in-depth molecular characterization of samples, capturing their morphology and RNA composition while retaining the spatial location (Fig. 1A). The gene expression profiles are usually available per spot, e.g., a 55µm tissue region (10x Genomics™, Visium) covering the whole transcriptome [3], or at a single-cell resolution but with a limited number of captured genes (CosMx NanoString™). More recent spatial transcriptomics technologies provide whole-transcriptome coverage along with higher resolution (e.g., 2µm, 10x Genomics™, Visium HD). Spatially aligning cell types by molecular phenotypes and morphology is important for understanding tissue-specific properties (e.g., neural organization in the brain [4]) in a physiological state and in the context of disease progression and treatment [5–8]. Nevertheless, spatial transcriptomics analysis demands manual annotation of multi-modal data, representing a laborious and resource-intensive process. Achieving reliable automation and overcoming limitations in cross-modal expertise will lead to more accurate annotations, offering a comprehensive, multi-modal perspective on biological mechanisms and interactions [9].

**Fig. 1.**
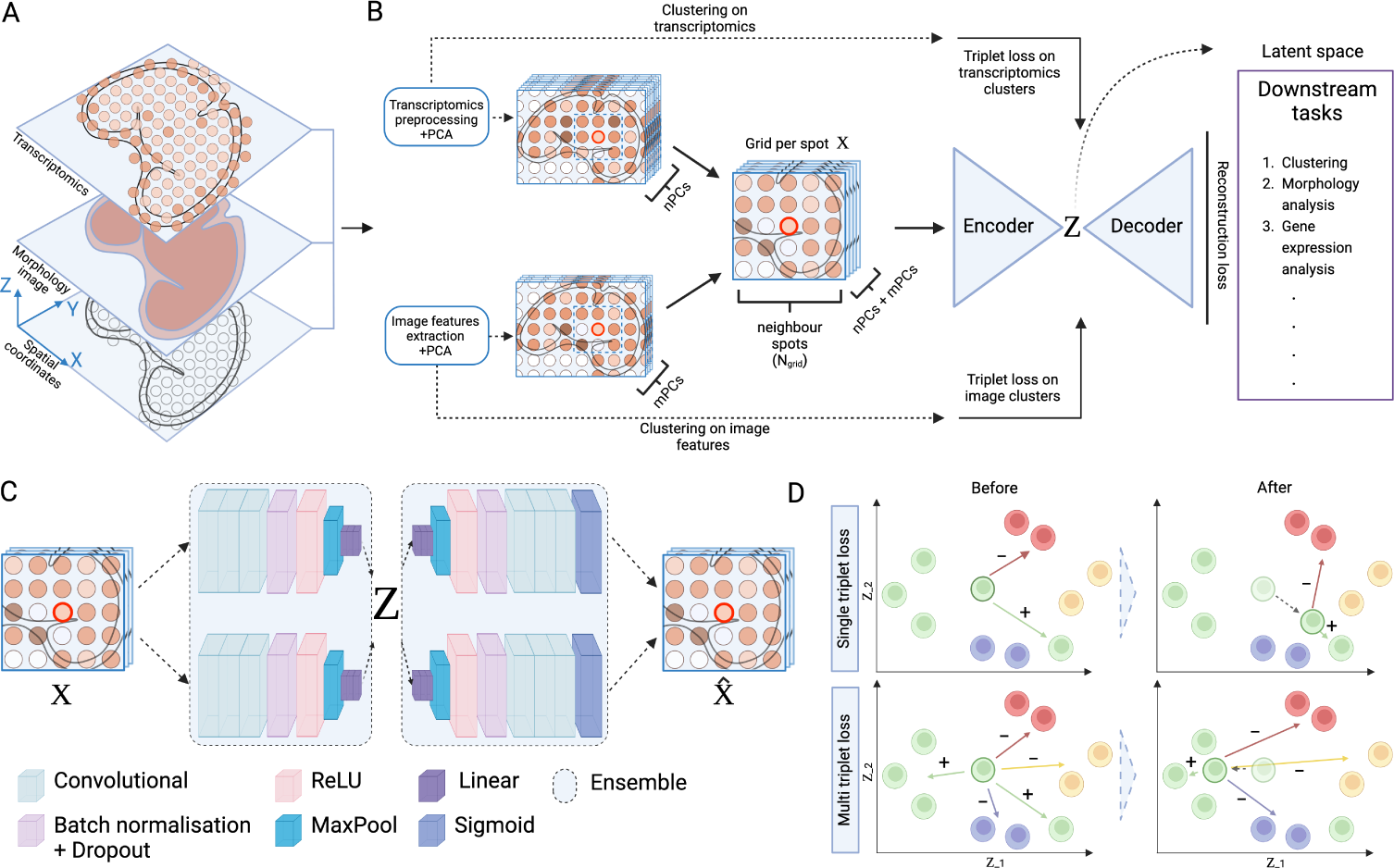
*AESTETIK* integrates spatial, transcriptomics, and morphology information to learn accurate spot representations. **A**: Spatial transcriptomics enables in-depth molecular characterization of samples on a morphology and RNA level while preserving spatial location. **B**: Workflow of AESTETIK. Initially, the transcriptomics and morphology spot representations are preprocessed. Next, a dimensionality reduction technique (e.g., PCA) is applied. Subsequently, the processed spot representations are clustered separately to acquire labels required for the multi-triplet loss. Afterwards, the modality-specific representations are fused through concatenation and the grid per spot is built. This is used as an input for the autoencoder. Lastly, the spatial-, transcriptomics-, and morphology-informed spot representations are obtained and used for downstream tasks such as clustering, morphology analysis, etc. **C**: *AESTETIK* relies on a convolutional encoder-decoder architecture to learn accurate spot representations from the spatial transcriptomics data. **D**: Employing a multi-triplet loss, instead of a single triplet loss adds extra positive and negative instances per class around the anchor point, improving the placement of the anchor in the latent space.

Despite recent progress, computational data analysis that integrates all available data modalities i.e., spatial information, transcriptomics, and morphology, remains challenging. Most existing methods either fall short in effectively integrating all modalities, especially those adapted from single-cell analysis or are computationally expensive [9]. For example, *BayesSpace* employs a Bayesian approach with a prior giving higher weight to physically close spots [10]; *MUSE* relies on a multi-view autoencoder to learn a latent space from transcriptomics and morphology [11]; *stLearn* quantifies morphological distance through histology image features and incorporates these distances with spatial neighbors to refine gene expression [12]. Furthermore, alternative methods suggested a different perspective on modeling the spatial transcriptomics data by employing graph neural networks (GNN) [13–15]. However, the expression profiles often suffer from biological variability (e.g., cellcycle stage) [16, 17] or technical noise [17, 18]. GNNs’ inherent susceptibility to noise can undermine their robustness and performance in downstream applications [19, 20]. Therefore, a new and reliable integration approach is needed to overcome the aforementioned challenges and improve spatial transcriptomics analysis, ensuring adaptability across spatial transcriptomics technologies.

To this end, we developed *AESTETIK*, a model that jointly integrates spatial, transcriptomics, and morphology information to learn accurate spot representations. We compared its performance against previous state-of-the-art methods on multiple datasets and widely adopted technology platforms: Brain tissue [21], breast cancer [22] and new and yet unreleased metastatic melanoma samples sequenced using Visium from 10x Genomics™; liver from normal and cancer patients using CosMx from NanoString™. We substantially improved the clustering accuracy across all datasets which yielded spatial domains with coherent expression and morphology. Through an ablation study, we showed the enhanced value of utilizing all available data modalities given the specifics of the analyzed tissue. Further, we validated the learned representation by identifying the main biological drivers and characterizing clusters based on morphology and cell-type composition.

## 2 Results

### 2.1 *AESTETIK* integrates spatial, transcriptomics, and morphology information

We introduce *AESTETIK* (**A**uto**E**ncoder for **S**patial **T**ranscriptomics **E**xpression with **T**opology and **I**mage **K**nowledge), a convolutional autoencoder model (Fig. 1B). It jointly integrates transcriptomics and morphology information at a spot level and topology at a neighborhood level to learn accurate spot representations that capture biological complexity. Firstly, we preprocess the transcriptomics profiles and apply principal component analysis (PCA) [23]. Simultaneously, the pre-trained on *Imagenet* [24] deep-learning model, *Inception v3* [25], is employed to extract morphology spot features, followed by PCA. After computing clusters separately to preserve the modality-specific structure, we concatenate the top principal components (PC) from both modalities. Next, we construct a square grid for each spot that includes spatially neighboring spots. This grid per spot, along with the precomputed clusters, serves as an input to *AESTETIK*. The model relies on a convolutional encoder-decoder architecture (Fig. 1C) to learn accurate spatial-, transcriptomics-, and morphology-informed spot representations. Ultimately, the learned representations can be leveraged for various downstream applications, including but not limited to clustering, gene expression, morphology, and pathway analysis.

The motivation for the grid construction is to form an image-like representation, with grid encoding for spatial neighborhood and channels for both transcriptomics and morphology modalities. We frame the machine-learning problem as image pattern recognition and compression, with convolutional autoencoders being the state-of-the-art architecture for addressing these challenges [26, 27]. The bottleneck layer serves as a constriction for information flow, forcing the model to capture the biological signal. Moreover, *AESTETIK*’s loss function (Eq. 4) is designed to optimize multiple objectives simultaneously by combining reconstruction loss for accurate latent representation and multi-triplet loss (Fig. 1D, Eq. 3) for structure preservation across modalities. This dual optimization ensures a comprehensive and informative representation of each data modality.

### 2.2 *AESTETIK* improves the identification of spatial domains

We benchmarked *AESTETIK* performance on multiple datasets with available ground truth annotations (Fig. 2A). In line with the methodology of [10–15], we adopted the Adjusted Rand Index (ARI) to measure the similarity between predicted cluster labels and ground truth, with the number of clusters set to match that in the ground truth. To avoid hyperparameter tuning on the samples used for testing, we introduced reversed leave-one-out cross-validation. More specifically, we used a single sample and its replicates to select hyperparameters to maximize the median ARI. Then, the optimal hyperparameters were applied to the remaining test samples. This process was iterated over all folds, and the resulting median ARI, along with the standard error, is reported (Fig. 2A).

**Fig. 2.**
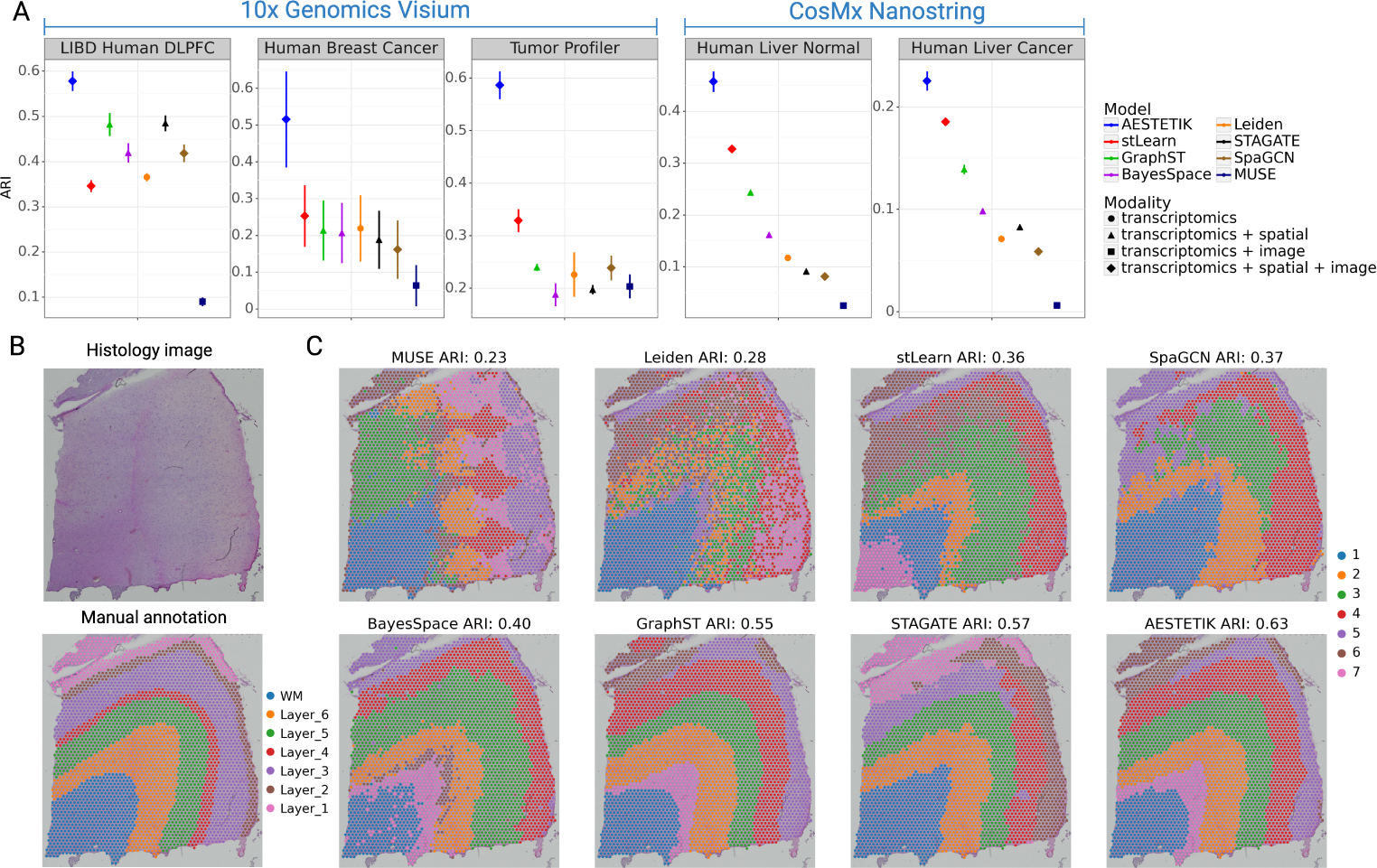
*AESTETIK* improves the identification of spatial domains with coherent expression and morphology. **A**: Benchmark of *AESTETIK* and previous state-of-the-art methods in spatial transcriptomics on 5 datasets across 2 technology platforms. The y-axis represents the ARI between the ground truth and the predicted labels. Models are ordered based on their relative rank across the datasets. The shape represents the modalities the model integrates. **B**: Histology image and manual annotation of slice 151676 from the LIBD human DLPFC dataset [21] and **C**: Comparison of cluster assignments for the same slice.

*AESTETIK* consistently yielded substantially improved cluster assignments closer to the ground truth annotations over previous state-of-the-art methods across all datasets (Fig. 2A). For example on the LIBD Human DLPFC dataset [21], *AESTETIK* achieved the highest ARI of 0.58*±*0.02, significantly surpassing the second best model - *GraphST* - by 21%. The LIBD Human DLPFC dataset comprises 12 tissue slices obtained from the dorsolateral prefrontal cortex (DLPFC) brain region, sequenced using Visium from 10x Genomics™, together with curated manual annotations based on brain cytoarchitecture and known marker genes (Fig. 2B). This improvement highlights the superior performance of *AESTETIK* in effectively integrating the spatial modality and generating accurate cluster assignments in structured brain tissue. *STAGATE* and *GraphST* demonstrated lower performance, achieving ARI of 0.48*±*0.02 and 0.48*±*0.03, respectively.

To qualitatively illustrate the cluster assignments, we compare them for slice 151676 (Fig. 2C), using the closest annotations to the ground truth across folds. *MUSE* (ARI 0.23), *Leiden* (ARI 0.28), *stLearn* (ARI 0.36) and *SpaGCN* (ARI 0.37) mixed the brain layers, accompanied by noise along the boundaries. *BayesSpace* (ARI 0.40) partitioned the white matter (WM) and layer 6 into multiple groups. While *GraphST* (ARI 0.55) and *STAGATE* (ARI 0.57) generated mostly well-defined clusters, layers 1, 2 and 3 were inconsistent. Notably, *AESTETIK* (ARI 0.63) identified the brain architecture, and its clusters displayed clearer definitions at the boundaries, leading to superior performance (Fig. 2C, S1).

Next, we investigate the methods’ performance on the Human Breast Cancer dataset, which comprises 5 tissue slices sequenced using Visium from 10x Genomics™ and annotated independently in two different labs [22]. This dataset presents unique challenges, primarily stemming from the considerable inter- and intra-sample heterogeneity, including variations in the cancer cell population. *AESTETIK* achieved the closest clusters to the ground truth labels with an ARI of 0.51*±*0.13 (Fig. 2A), indicating a two-fold increase over the second best model, *stLearn* (ARI 0.25*±*0.08). Despite wide standard error intervals observed in all models, *AESTETIK* exhibited heightened robustness, surpassing the challenges posed by the complexity of breast cancer tissue (Fig. S2).

### 2.3 *AESTETIK* effectively incorporates the morphology modality

We introduce a new and yet unreleased spatial transcriptomics dataset with 9 distinct tissue regions sequenced using Visium from 10x Genomics™ from the Tumor Profiler study [28]. Each region has a replicate resulting in 18 samples of size 6.5 *×* 6.5mm^2^, with data including 10x Genomics™ *Space Ranger v3.0.0* outputs and a corresponding H&E image scanned at a high resolution of 0.3µm/pixel. The tissue regions originate from 7 patients with metastatic melanoma each characterized by one of the following immune subtypes: immune desert, immune excluded, or inflamed. The ground truth annotations were obtained using histopathology software (HALO AI™ (Indica Labs, Corrales, NM, USA)), classifying the spots into one of the following categories: tumor, stroma, normal lymphoid, and blood/necrosis. Following this, a pathologist manually reviewed the model predictions (Fig. 3A). We consider this dataset to be a valuable reference benchmark for evaluating the performance of spatial transcriptomics models, particularly in terms of their ability to integrate morphology effectively.

**Fig. 3.**
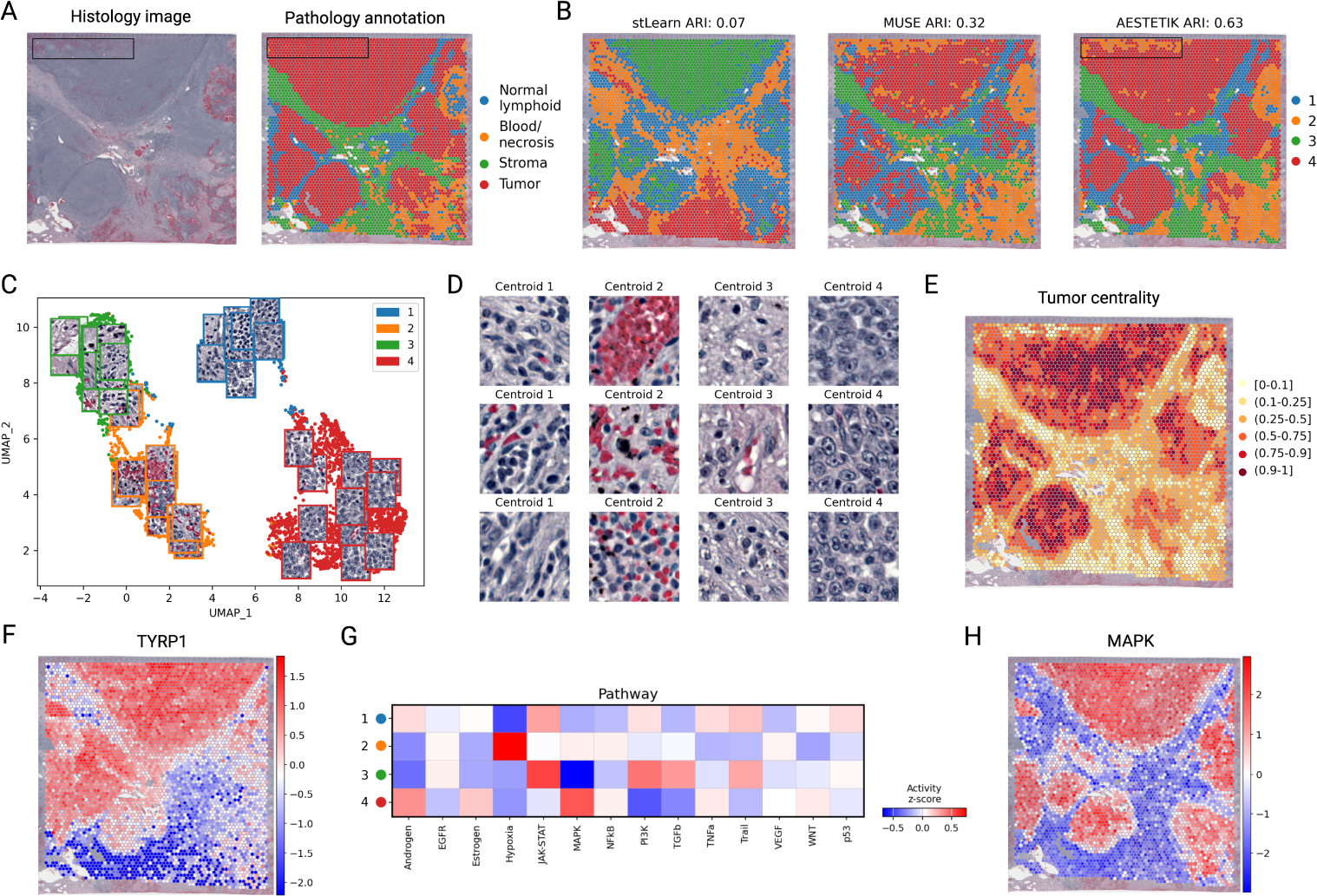
*AESTETIK* effectively incorporates the morphology modality revealing biologically relevant spatial organization of cancer tissue. **A**: Histology image and pathology annotation of slice MACEGEJ-2-2 from the Tumor Profiler dataset. **B**: Comparison of cluster assignments for slice MACEGEJ-2-2. **C**: *UMAP* plot of the *AESTETIK*’s latent space with randomly sampled spot images. **D**: Most representative cluster spots based on the obtained representations. **E**: Euclidean distance in latent space of each spot to the tumor centroid plotted in spatial space. Most representative spots are located in the middle of the tumor formations. **F**: Spatial marker gene expression of TYRP1. **G**: Pathway analysis of the identified clusters using *decoupler* [29]. **H** Spatial activation of MAPK pathway.

On this dataset, *AESTETIK* achieved a 79% increase in ARI (0.59*±*0.03) over previous state-of-the-art methods, demonstrating effective use of the morphology modality (Fig. 2A). While both *stLearn* (ARI 0.33*±*0.02) and *MUSE* (ARI 0.20*±*0.02) use the same pre-trained *Inception v3* [25] for extracting morphology features, they fall short in effectively leveraging this information (Fig. 3B). On the other hand, *AESTETIK* not only produced accurate cluster assignments (Fig. 3B), but also identified a hemorrhage region in the upper left of the H&E image, that was overlooked during annotation (black box in Fig. 3A, B, S3).

Next, we qualitatively explore the latent representations and the identified spatial domains by focusing on slice MACEGEJ-2-2 (Fig. 3A). We visualized the latent space using *UMAP* [30] with randomly sampled morphology spot representations (Fig. 3C).

We observe an aggregation of tumor spots (cluster 4) on the bottom-right side, showcasing similarities in their characteristics. On the lower left side, there are areas with blood and necrosis. While clusters 1 and 3, representing normal lymphoid tissue and stromal cells, are positioned in the upper part, a closer inspection reveals discernible differences in their underlying structures. Besides, for enhanced explainability of the spot representations, we selected the most representative spots per cluster (Fig. 3D). Visually, tumor cells within cluster 4 exhibit distinct characteristics; they appear significantly larger, displaying irregular shapes, and possessing enlarged nuclei. Stromal cells (cluster 3) have an elongated morphology and are noticeably more scattered [31] compared to the normal lymphoid cells (cluster 1), which are generally smaller and denser [32].

Furthermore, to illustrate the effect of encoding spatial information in latent space, we computed the Euclidean distance of each spot to the tumor centroid and visualized it spatially (Fig. 3E). While stromal cells are the furthest, we observed that tumor cells close in latent space are clustered spatially, with the most representative spots located in the middle of the tumor formations.

To provide additional insights, we found TYRP1 and TKTL1 among the top tumor marker genes in the tumor cluster, confirming the model predictions for the spatially resolved identification of melanoma cells (Fig. 3F, S4). TYRP1 gene is involved in melanocyte pigmentation, associated with melanoma progression and is a target for oncological immunotherapy [33–35]. The second highly upregulated gene, TKTL1, is implicated in the progression of melanoma and contributes to the increased invasion of melanoma cells [36]. Further, we performed a pathway analysis of the tumor clusters using *decoupler* [29] (Fig. 3G) which revealed increased activity of the MAPK pathway in the cancer cluster (Fig. 3H), known for promoting cell proliferation, invasion, metastasis, migration, survival, and angiogenesis [37–39]. Furthermore, we observed that hypoxia signatures were predominant in areas of necrosis and hemorrhage and JAK-STAT inflammatory signaling was predominant in the cancer microenvironment clusters 1 and 3. These pathway-level analyses underline the robust associations of the spatially resolved clustering results achieved by *AESTETIK* and support interpretation in the context of the underlying biology.

### 2.4 *AESTETIK* improves cluster assignment in single-cell spatial transcriptomics

CosMx NanoString™ released a liver dataset with single-cell resolution, encompassing two tissue regions from normal and cancer patients and capturing 1000 genes. It offers valuable insights into liver biology and cancer characteristics. More specifically, using these datasets, we assess the models’ effectiveness at single-cell resolution by comparing the clusters they produce with the cell types reported by NanoString™. In both normal and cancer liver tissue, *AESTETIK* exhibited outstanding performance, substantially outperforming the other models by 39% and 21%, with ARI of 0.46*±*0.02 and 0.23*±*0.00, respectively (Fig. 2A, S5, S5). The second best model, *stLearn*, attained a score of 0.33*±*0.00 and 0.19*±*0.00, followed by *GraphST* with 0.24*±*0.00 and 0.14*±*0.00. Overall, the clustering accuracy on the cancer tissue is lower compared to the normal sample. However, the relative trend in the ranking of the models remained consistent.

### 2.5 Joint integration of multi-modal data enhances computational analysis

To pinpoint the benefit of the spatial modality in the LIBD Human DLPFC and Tumor Profiler datasets, we systematically varied the grid’s window size, ranging from 1 (w/o spatial information) to 11, and measured the change in ARI (Fig. 4A). The grid size determines the number of spatially adjacent spots to consider. Local spatial information proved important, preserving local details and spot-to-spot variability. However, incorporating a more extensive global context through a larger window size (e.g., 11) introduced noise and hampered performance, which was likely due to signal over-smoothing and the loss of spot-specific details.

**Fig. 4.**
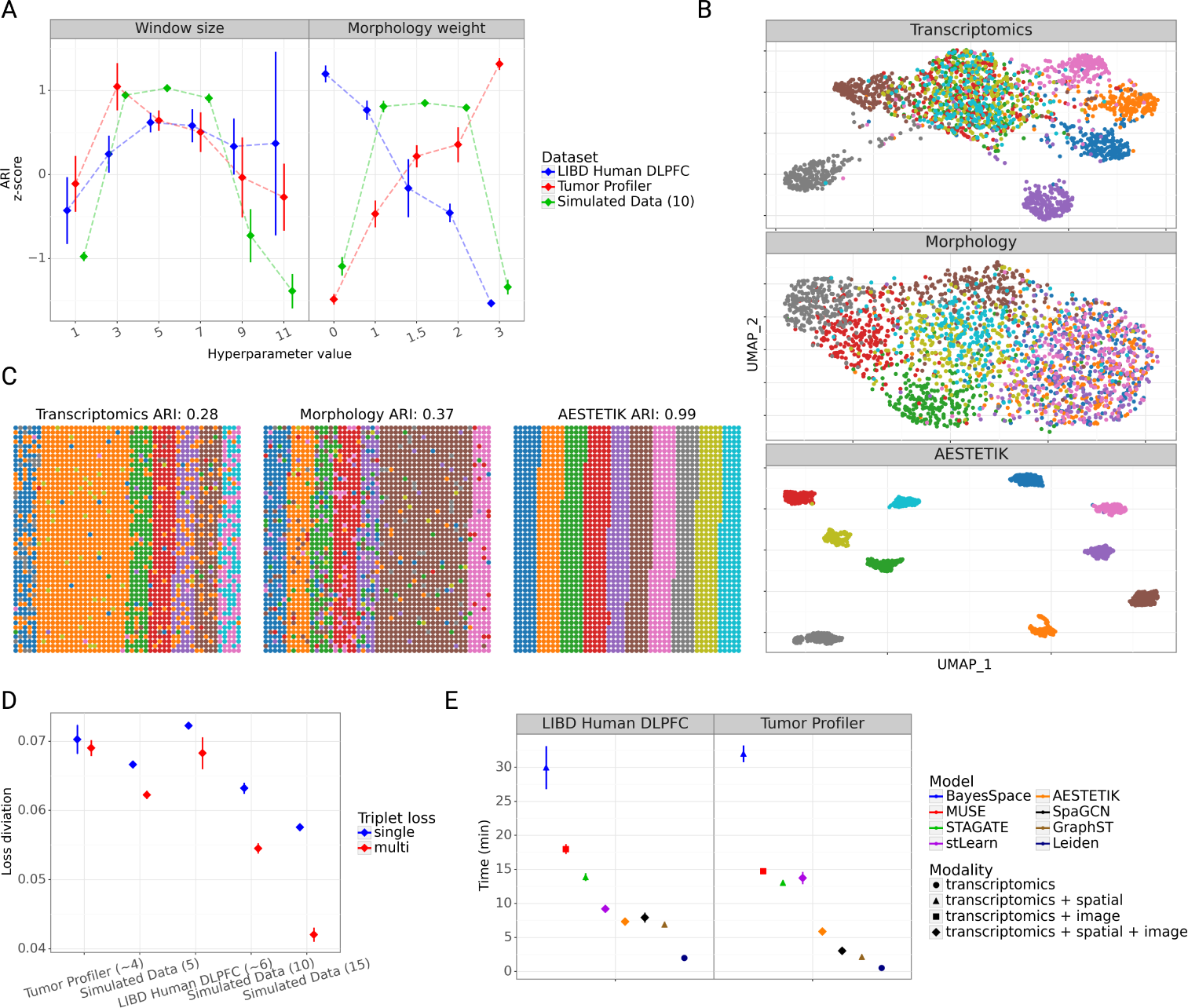
Joint integration of multi-modal data enhances computational analysis. **A**: Ablation study on the influence of window size and morphology weight on the ARI. The y-axis represents the ARI, normalized by dataset. **B**: *UMAP* visualization of single (transcriptomics, morphology) and combined (*AESTETIK*) modality representations on a simulated tissue slice, colored by the 10 ground truth annotations. **C**: Cluster assignments based on only a single modality (transcriptomics, morphology) and *AESTETIK*’s joint representations. **D**: Comparing the stability of the single and multi-triplet loss on the loss function of *AESTETIK* across datasets. The number, following the dataset name, is the median number of clusters present. The y-axis represents the standard deviation computed on the loss difference over successive training epochs. **E**: Runtime for the evaluated clustering methods. The y-axis represents the time in minutes. Models are ordered based on their relative rank across the datasets.

Similarly, we studied the contribution of each modality by varying the morphology weight (0 - no morphology; 1.5 - equal weight between transcriptomics and morphology; 3 - only morphology). As expected, we found that the transcriptomics modality in the brain dataset is informative given the provided ground truth annotations, relying on known cytoarchitecture and marker genes [21]. In contrast, the ground truth for the Tumor Profiler, derived from histopathology software, was morphology driven (Fig. 4A). Furthermore, to underscore the significance of methods incorporating all data modalities, we present a scenario illustrating the necessity of both modalities to reveal ground truth annotations (Fig. 4B). Following the approach of [11], we simulated data where both modalities are essential for accurate cluster identification. Our ablation study demonstrated that the optimal ARI was achieved when accounting for both modalities, thus emphasizing the critical significance of multi-modal data integration (Fig. 4C).

Additionally, the multi-triplet loss demonstrated an enhancement in loss stability during training compared to the single triplet loss (Fig. 4D). The refined positioning of clusters in latent space, considering multiple positive and negative spots, becomes crucial, especially when dealing with datasets containing numerous clusters. Lastly, the runtime per tissue slice for our model, incorporating all three modalities (*∼*8 min), was either lower or comparable to that of other models (Fig. 4E). For example, *BayesSpace*, *MUSE*, and *STAGATE*, incorporating only two modalities, required *∼*28 min, *∼*17 min, and *∼*13 min, respectively. Moreover, we demonstrate that *AESTETIK* is well-suited for analyzing large spatial transcriptomics datasets, scaling to millions of spots (Fig. S7).

## 3 Discussion

In this work, we propose *AESTETIK*, a method that jointly integrates spatial, transcriptomics, and morphology information to learn accurate spot representations. Our results consistently showed superior performance to state-of-the-art methods across structured tissues (e.g., brain) and cancer tissues with heterogeneous cell populations (e.g., breast, melanoma, liver) across widely adopted spatial transcriptomics technologies (e.g., 10x Genomics™, Visium, CosMx NanoString™). We systematically demonstrated the significance of jointly integrating multi-modal data to improve spatial transcriptomics analysis and yield more precise spot annotations.

This improvement in spot representation resulted from modeling the spatial transcriptomics modalities as a grid encoding the spatial spot neighborhood and channels as transcriptomics and morphology modalities. Our approach framed the machine-learning problem as image pattern recognition and compression, where convolution filters jointly learn the importance of neighboring spots and channels. This proved beneficial in both structured and heterogeneous tissues. In contrast, the GNNs, employed by *SpaGCN, GraphST* and *STAGATE* demonstrated variations in their performance relative to the other methods across tissue types and spatial transcriptomics technologies. This could be attributed to the inherent susceptibility of GNNs to noise [19, 20]. The graph structure ensures connectivity among neighboring spots, which is useful in structured tissues (e.g., brain) with coherent spatial patterns. However, it presents challenges in samples of lower sequencing quality or tissues with higher heterogeneity (e.g., cancer cell populations), where noise, introduced through node perturbations and edge alterations, might affect the graph structure. Consequently, this undermines the robustness and performance of current GNNs in downstream applications.

Further, in our ablation study on the brain dataset, we quantitatively demonstrated the significance of the spatial modality in identifying the brain layer structure. We discovered that a relatively small grid’s window size (5-7) sufficiently captures the desired spatial signal. Opting for a larger neighborhood (e.g., window size 11) offers no extra value. Unlike the global tissue context, the local environment better preserves spot-specific signals and nearby variability. Ultimately, our ablation results underscore **1)** the importance of jointly integrating the available spatial transcriptomics data modalities for accurate spot representation, and **2)** the necessity for external knowledge to prioritize the signal of interest, depending on the particular research question at hand.

Several paths to further improve model accuracy appear promising. **1)** We employed the pre-trained *Inception v3* [25] to extract morphology features. However, adopting a model tailored to a specific task (e.g., cell nuclei segmentation and classification) would likely yield more informative spot features, potentially leading to improved overall performance. **2)** *AESTETIK* randomly selects the positive and negative pairs for each anchor point during training. We believe this process can be improved by utilizing a *smarter* strategy for triplet mining, which would eventually improve the performance, and robustness to noise.

In the future, *AESTETIK* could be effectively applied to fine-map cell populations in spatial transcriptomics datasets [40–42], to systematically analyze the interplay between different modalities by varying their contribution and to gain a multi-modal understanding of key biological processes. To foster these downstream applications, we have released the code for *AESTETIK* along with examples demonstrating its usage. Moreover, we anticipate that upon its release, the 10x Genomics™, Visium dataset from the Tumor Profiler study will serve as a valuable reference benchmark for assessing spatial transcriptomics model performance and explainability. Thus, we hope that our model, together with this dataset, will stimulate further improvements in computational spatial transcriptomics analysis.

## 4 Methods & Materials

### 4.1 Data preprocessing

*AESTETIK* takes in spatial, transcriptomics, and morphology information. We apply the same preprocessing pipeline across datasets and sequencing technologies. For simplicity, we refer to both spot and cell as a spot *(a single spot can contain 1 cell)*.

#### 4.1.1 Transcriptomics modality

Starting with raw counts, genes expressed in fewer than 10 spots are removed. Then, the *scanpy* function *highly variable genes* computes normalized variance in *Seurat v3* style, removing genes with variance below 1 [23, 43]. Each spot undergoes normalization by total counts over all genes, followed by log1p transformation and scaling. Subsequently, PCA is applied to the preprocessed counts, extracting the first 15 PCs [10].

#### 4.1.2 Morphology modality

The raw RGB image for each tissue slice is divided into tails, each representing a spot and its defined neighborhood based on the spot diameter. Following the default preprocessing steps of *Inception v3* [25], the tiles are resized to 299, their center is cropped and the RGB channels are normalized. Morphology features are then extracted from the last network layer (with 2,048 dimensions) of the pre-trained on *Imagenet* [24] deep-learning model *Inception v3*. Finally, PCA reduces the feature dimension from 2,048 to 15.

#### 4.1.3 Grid construction

To begin, each spot is represented by two vectors containing the first *n_d_*_1_ and *n_d_*_2_ PCs obtained via PCA from the preprocessed transcriptomics and morphology modalities, along with their spatial coordinates. For simplicity, we assume *n_d_*_1_ = *n_d_*_2_ = *n_pca_*, but the following workflow holds also for *n_d_*_1_ ≠ *n_d_*_2_. These vectors are concatenated and scaled in the range [0, 1]. Then, a square grid for each spot is constructed with the number of spatial neighbors, *N_grid_*, chosen as an odd number to ensure the center position of the selected spot in the window. This results in a tensor of size:

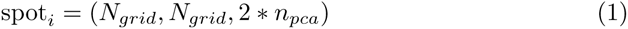

which can be interpreted as *N_grid_* × *N_grid_*image with 2 *∗ n_pca_*channels. For missing or located on the borders spots, we apply padding by taking the median expression over each channel in spot*_i_*.

#### 4.1.4 Clustering

The default clustering algorithm is *Bayesian Gaussian Mixture* with a diagonal covariance matrix from the *sklearn* package, but we also support *K-Means, Leiden*, and *Louvain*. Once the cluster labels are obtained, an additional preprocessing step can be applied. A K-Neighbors Classifier is fitted using spatial coordinates and the already obtained clusters to refine the cluster assignments in spatial space through majority voting.

### 4.2 Model architecture

*AESTETIK* utilizes a convolutional deep-learning autoencoder with a standard encoder-decoder architecture and a bottleneck layer. The encoder comprises a convolutional layer, max-pooling layer, batch normalization, *ReLU* activation, and linear layer. Default hyperparameters include 64 convolutional kernels (size 7), dropout (*p* = 0.3), max-pooling (stride 3), and a linear layer (size 16). The decoder follows a mirrored architecture, concluding with a sigmoid function to constrain output values in the range [0, 1]. *AESTETIK* is a *Python* package implemented in *PyTorch*.

#### 4.2.1 Autoencoder ensemble

To improve the network stability, we employ ensemble architectures for both the encoder and decoder, utilizing random *LeCun* [44] initialization. The ensemble’s output is determined by taking the median over predictions. We train an ensemble with 3 encoders and decoders. The final representation is computed by dropout sampling 1,000 times and taking the median value.

#### 4.2.2 Reconstruction loss

We employ a reconstruction loss to ensure that the latent space effectively captures the biological complexity of the morphology and transcriptomics modalities. We define it as:

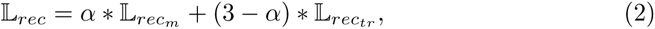

where *α ∈* [0,3] is a hyperparameter for the morphology weight. 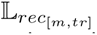 is the standard L1 reconstruction loss with *m* for morphology and *tr* for transcriptomics modality. We use L1 loss due to the input-output range being [0, 1].

#### 4.2.3 Multi-triplet loss

We apply triplet loss to preserve the structure across modalities. Its primary objective is to learn a spot representation in which similar instances are closer together, while dissimilar instances are farther apart. Define an anchor point *A* with label *l_i_*, then we draw at random a positive point *P* with label *l_i_* and a negative point *N* with label *l_j_* such as *l_i_* ≠ *l_j_*, then the single triplet loss is defined as:

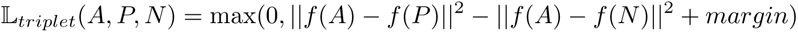

In spatial transcriptomics, multiple classes and high noise ratios are typical. Using just one positive and negative point can lead to unstable representation and increase the training time due to alternations. To improve the spot representation robustness, we propose the multi-triplet loss motivated by [45]. Let *L* be the number of unique labels in the dataset. Define an anchor point *A* with label *l_i_*, then we draw, with replacement, *L −* 1 positive points *{P*_1_*, P*_2_*, …, P_L−_*_1_*}* with label *l_i_*. Additionally, for each label *l_j_* where *j* ≠ *i*, we draw a single negative point, resulting in *{N*_1_*, N*_2_*, …, N_L−_*_1_*}*. Then the multi-triplet loss for a single modality can be defined as:

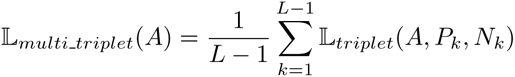

which when extended to all spots:

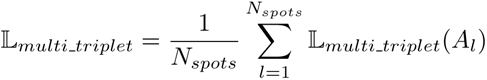

The modality-weighted multi-triplet loss is defined as:

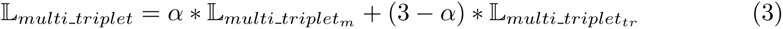

with *α* defined as in equation 2.

#### 4.2.4 Loss function

The overall loss function for training combines reconstruction loss to ensure accurate latent representation and multi-triplet loss for preserving structure across modalities. Formally, it is defined as:

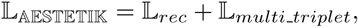

which can be rewritten as:

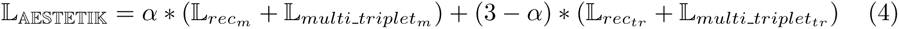

#### 4.2.5 Training details

The model is trained for 100 epochs using *Adam* [46] with a weight decay of 1e-6, a learning rate of 1e-3, and a batch size corresponding to the number of spots in a tissue slice. The run time is approximately 8 minutes on a GPU, with inference time under a minute. Computational data analysis was performed at Leonhard Med (https://sis.id.ethz.ch/services/sensitiveresearchdata/) secure trusted research environment at ETH Zurich.

#### 4.2.6 Evaluation

We propose reversed leave-one-out cross-validation for model evaluation to avoid hyperparameter tuning on test samples. We utilize a single sample and its replicates to select hyperparameters through a grid search, aiming to maximize the median ARI. Subsequently, the optimal hyperparameters are applied to the remaining test samples. For state-of-the-art methods, we consider hyperparameter values suggested by the authors, as well as those discussed in the corresponding paper. Hyperparameter values are provided in the supplement. To ensure comparable conditions across models, the number of clusters is pre-defined based on the provided ground truth. Performance is assessed using the ARI between ground-truth labels and cluster assignments. We bootstrap 10,000 times from the median ARI across the test folds and report the resulting median ARI and its standard error. For most datasets, we generated all possible sample combinations (folds). However, for the larger CosMx NanoString™ Liver dataset we use each FOV to select the hyperparameters, but we evaluate on randomly selected 20 FOVs, not adjacent to the FOVs used for optimization.

#### 4.2.7 Ablation study

In the ablation study on *AESTETIK*, we adhere to the procedure described in 4.2.6, where we fix the value for the hyperparameter of interest and assess its impact on the ARI. To ensure comparability across datasets, we compute an ARI z-score.

### 4.3 Downstream applications

#### 4.3.1 Marker genes

For marker genes, we employ the *rank genes groups* function from *scanpy* using the Wilcoxon signed-rank test. Significant marker genes (adjusted p-value *<* 0.05) are selected and sorted by their average log-fold change. The top 15 marker genes per cluster are reported.

#### 4.3.2 Pathway analysis

For pathway analysis, we utilize the multivariate linear model from the *decoupler* package [29] to compute regulatory pathway activities from the *PROGENy* database [47].

#### 4.3.3 Cluster centroids in latent space

To determine the centroid for each cluster in the latent space, we employed a method minimizing the sum of Euclidean distances among all samples within that class. Subsequently, we computed the top N spots near each cluster centroid.

### 4.4 Data availability

The LIBD Human DLPFC dataset is available at https://github.com/LieberInstitute/HumanPilot and http://research.libd.org/spatialLIBD; Human Breast Cancer - Zenodo https://doi.org/10.5281/zenodo.4739739, Human Liver Normal and Cancer - https://nanostring.com/products/cosmx-spatial-molecular-imager/human-liver-rna-ffpe-dataset/. The metastatic melanoma dataset with 18 tissue slices from Tumor Profiler samples sequenced using Visium from 10x Genomics™ will be made available upon acceptance of publication.

### 4.5 Code availability

The open-source implementation of *AESTETIK* along with a tutorial is available at:

- www.github.com/ratschlab/aestetik

The Snakemake pipeline for reproducing the results is available at:

- www.github.com/ratschlab/st-rep

## Acknowledgments

This work was supported by the Swiss Federal Institutes of Technology (strategic focus area of personalized health and related technologies; 2021–367). The 10x spatial transcriptomics sequencing of the Tumor Profiler samples was made possible through a technology access program by 10x Genomics™, with special acknowledgments to Jacob Stern, James Chell, Rudi Schläfli, Laura Lipka, Mario Werner, Nikhil Rao, and Scott Brouilette for their invaluable contributions. The Tumor Profiler study was supported by a public-private partnership involving Roche Holding AG, ETH Zurich, University of Zurich, University Hospital Zurich, and University Hospital Basel. V.H.K. gratefully acknowledges additional funding by the Promedica Foundation (F-87701-41-01).

## TUMOR PROFILER CONSORTIUM

Rudolf Aebersold^5^, Melike Ak^33^, Faisal S Al-Quaddoomi^12,22^, Silvana I Albert^10^, Jonas Albinus^10^, Ilaria Alborelli^29^, Sonali Andani^9,22,31,36^, Per-Olof Attinger^14^, Marina Bacac^21^, Daniel Baumhoer^29^, Beatrice Beck-Schimmer^44^, Niko Beerenwinkel^7,22^, Christian Beisel^7^, Lara Bernasconi^32^, Anne Bertolini^12,22^, Bernd Bodenmiller^11,40^, Ximena Bonilla^9^, Lars Bosshard^12,22^, Byron Calgua^29^, Ruben Casanova^40^, Stéphane Chevrier^40^, Natalia Chicherova^12,22^, Ricardo Coelho^23^, Maya D’Costa^13^, Esther Danenberg^42^, Natalie R Davidson^9^, Monica-Andreea Dragan^7^, Reinhard Dummer^33^, Stefanie Engler^40^, Martin Erkens^19^, Katja Eschbach^7^, Cinzia Esposito^42^, André Fedier^23^, Pedro F Ferreira^7^, Joanna Ficek-Pascual^1,9,16,22,31^, Anja L Frei^36^, Bruno Frey^18^, Sandra Goetze^10^, Linda Grob^12,22^, Gabriele Gut^42^, Detlef Günther^8^, Pirmin Haeuptle^3^, Viola Heinzelmann-Schwarz^23,28^, Sylvia Herter^21^, Rene Holtackers^42^, Tamara Huesser^21^, Alexander Immer^9,17^, Anja Irmisch^33^, Francis Jacob^23^, Andrea Jacobs^40^, Tim M Jaeger^14^, Katharina Jahn^7^, Alva R James^9,22,31^, Philip M Jermann^29^, André Kahles^9,22,31^, Abdullah Kahraman^22,36^, Viktor H Koelzer^36,41^, Werner Kuebler^30^, Jack Kuipers^7,22^, Christian P Kunze^27^, Christian Kurzeder^26^, Kjong-Van Lehmann^2,4,9,15^, Mitchell Levesque^33^, Ulrike Lischetti^23^, Flavio C Lombardo^23^, Sebastian Lugert^13^, Gerd Maass^18^, Markus G Manz^35^, Philipp Markolin^9^, Martin Mehnert^10^, Julien Mena^5^, Julian M Metzler^34^, Nicola Miglino^35,41^, Emanuela S Milani^10^, Holger Moch^36^, Simone Muenst^29^, Riccardo Murri^43^, Charlotte KY Ng^29,39^, Stefan Nicolet^29^, Marta Nowak^36^, Monica Nunez Lopez^23^, Patrick GA Pedrioli^6^, Lucas Pelkmans^42^, Salvatore Piscuoglio^23,29^, Michael Prummer^12,22^, Prélot, Laurie^9,22,31^, Natalie Rimmer^23^, Mathilde Ritter^23^, Christian Rommel^19^, María L Rosano-González^12,22^, Gunnar Rätsch^1,6,9,22,31^, Natascha Santacroce^7^, Jacobo Sarabia del Castillo^42^, Ramona Schlenker^20^, Petra C Schwalie^19^, Severin Schwan^14^, Tobias Schär^7^, Gabriela Senti^32^, Wenguang Shao^10^, Franziska Singer^12,22^, Sujana Sivapatham^40^, Berend Snijder^5,22^, Bettina Sobottka^36^, Vipin T Sreedharan^12,22^, Stefan Stark^9,22,31^, Daniel J Stekhoven^12,22^, Tanmay Tanna^7,9^, Alexandre PA Theocharides^35^, Tinu M Thomas^9,22,31^, Markus Tolnay^29^, Vinko Tosevski^21^, Nora C Toussaint^12,22^, Mustafa A Tuncel^7,22^, Marina Tusup^33^, Audrey Van Drogen^10^, Marcus Vetter^25^, Tatjana Vlajnic^29^, Sandra Weber^32^, Walter P Weber^24^, Rebekka Wegmann^5^, Michael Weller^38^, Fabian Wendt^10^, Norbert Wey^36^, Andreas Wicki^35,41^, Mattheus HE Wildschut^5,35^, Bernd Wollscheid^10^, Shuqing Yu^12,22^, Johanna Ziegler^33^, Marc Zimmermann^9^, Martin Zoche^36^, Gregor Zuend^37^

^1^AI Center at ETH Zurich, Andreasstrasse 5, 8092 Zurich, Switzerland, ^2^Cancer Research Center Cologne-Essen, University Hospital Cologne, Cologne, Germany, ^3^Cantonal Hospital Baselland, Medical University Clinic, Rheinstrasse 26, 4410 Liestal, Switzerland, ^4^Center for Integrated Oncology Aachen (CIO-A), Aachen, Germany, ^5^ETH Zurich, Department of Biology, Institute of Molecular Systems Biology, Otto-Stern-Weg 3, 8093 Zurich, Switzerland, ^6^ETH Zurich, Department of Biology, Wolfgang-Pauli-Strasse 27, 8093 Zurich, Switzerland, ^7^ETH Zurich, Department of Biosystems Science and Engineering, Mattenstrasse 26, 4058 Basel, Switzerland, ^8^ETH Zurich, Department of Chemistry and Applied Biosciences, Vladimir-Prelog-Weg 1-5/10, 8093 Zurich, Switzerland, ^9^ETH Zurich, Department of Computer Science, Institute of Machine Learning, Universitätstrasse 6, 8092 Zurich, Switzerland, ^10^ETH Zurich, Department of Health Sciences and Technology, Otto-Stern-Weg 3, 8093 Zurich, Switzerland, ^11^ETH Zurich, Institute of Molecular Health Sciences, Otto-Stern-Weg 7, 8093 Zurich, Switzerland, ^12^ETH Zurich, NEXUS Personalized Health Technologies, Wagistrasse 18, 8952 Zurich, Switzerland, ^13^F. Hoffmann-La Roche Ltd, Grenzacherstrasse 124, 4070 Basel, Switzerland, ^14^F. Hoffmann-La Roche Ltd, Grenzacherstrasse 124, 4070 Basel, Switzerland, ^15^Joint Research Center Computational Biomedicine, University Hospital RWTH Aachen, Aachen, Germany, ^16^Life Science Zurich Graduate School, Biomedicine PhD Program, Winterthurerstrasse 190, 8057 Zurich, Switzerland, ^17^Max Planck ETH Center for Learning Systems, ^18^Roche Diagnostics GmbH, Nonnenwald 2, 82377 Penzberg, Germany, ^19^Roche Pharmaceutical Research and Early Development, Roche Innovation Center Basel, Grenzacherstrasse 124, 4070 Basel, Switzerland, ^20^Roche Pharmaceutical Research and Early Development, Roche Innovation Center Munich, Roche Diagnostics GmbH, Nonnenwald 2, 82377 Penzberg, Germany, ^21^Roche Pharmaceutical Research and Early Development, Roche Innovation Center Zurich, Wagistrasse 10, 8952 Schlieren, Switzerland, ^22^SIB Swiss Institute of Bioinformatics, Lausanne, Switzerland, ^23^University Hospital Basel and University of Basel, Department of Biomedicine, Hebelstrasse 20, 4031 Basel, Switzerland, ^24^University Hospital Basel and University of Basel, Department of Surgery, Brustzentrum, Spitalstrasse 21, 4031 Basel, Switzerland, ^25^University Hospital Basel, Brustzentrum & Tumorzentrum, Petersgraben 4, 4031 Basel, Switzerland, ^26^University Hospital Basel, Brustzentrum, Spitalstrasse 21, 4031 Basel, Switzerland, ^27^University Hospital Basel, Department of Information- and Communication Technology, Spitalstrasse 26, 4031 Basel, Switzerland, ^28^University Hospital Basel, Gynecological Cancer Center, Spitalstrasse 21, 4031 Basel, Switzerland, ^29^University Hospital Basel, Institute of Medical Genetics and Pathology, Schönbeinstrasse 40, 4031 Basel, Switzerland, ^30^University Hospital Basel, Spitalstrasse 21/Petersgraben 4, 4031 Basel, Switzerland, ^31^University Hospital Zurich, Biomedical Informatics, Schmelzbergstrasse 26, 8006 Zurich, Switzerland, ^32^University Hospital Zurich, Clinical Trials Center, Ramistrasse 100, 8091 Zurich, Switzerland, ^33^University Hospital Zurich, Department of Dermatology, Gloriastrasse 31, 8091 Zurich, Switzerland, ^34^University Hospital Zurich, Department of Gynecology, Frauenklinikstrasse 10, 8091 Zurich, Switzerland, ^35^University Hospital Zurich, Department of Medical Oncology and Hematology, Rämistrasse 100, 8091 Zurich, Switzerland, ^36^University Hospital Zurich, Department of Pathology and Molecular Pathology, Schmelzbergstrasse 12, 8091 Zurich, Switzerland, ^37^University Hospital Zurich, Ramistrasse 100, 8091 Zurich, Switzerland, ^38^University Hospital and University of Zurich, Department of Neurology, Frauenklinikstrasse 26, 8091 Zurich, Switzerland, ^39^University of Bern, Department of BioMedical Research, Murtenstrasse 35, 3008 Bern, Switzerland, ^40^University of Zurich, Department of Quantitative Biomedicine, Winterthurerstrasse 190, 8057 Zurich, Switzerland, ^41^University of Zurich, Faculty of Medicine, Zurich, Switzerland, ^42^University of Zurich, Institute of Molecular Life Sciences, Winterthurerstrasse 190, 8057 Zurich, Switzerland, ^43^University of Zurich, Services and Support for Science IT, Winterthurerstrasse 190, 8057 Zurich, Switzerland, ^44^University of Zurich, VP Medicine, Künstlergasse 15, 8001 Zurich, Switzerland

## Consent for publication

This manuscript has been seen and approved by all listed authors. The figures were created using BioRender.com and exported under a paid subscription.

