## Supplementary Information for "Representation learning for multi-modal spatially resolved transcriptomics data"

#### 1 Comparison with other methods

For comparing other spatial transcriptomics methods, we applied the suggested preprocessing pipelines outlined in the corresponding papers and tutorials. A summary can be found here:

- *BayesSpace* [1]: Raw counts are normalized, log1p transformed, the 2000 most variable genes are selected and PCA is applied to select the first 15 PCs.
- *stLearn* [2]: Raw counts are normalized, log1p transformed, and PCA is applied to select the first 15 PCs. For morphology features *Inception v3* is used.
- *GraphST* [3]: The first 3000 most variable genes are selected in *Seurat v3* style. Then the raw counts are normalized, log1p transformed and scaled.
- *STAGATE* [4]: The first 3000 most variable genes are selected in *Seurat v3* style. Then the raw counts are normalized and log1p transformed.
- *MUSE* [5]: The first 500 most variable genes are selected in *Seurat v3* style. Then the raw counts are normalized, log1p transformed and PCA is applied to select the first 15 PCs. For morphology features *Inception v3* is used.
- *SpaGCN* [6]: The MT and ERCC genes are removed along with genes expressed in less than 3 cells. Then the raw counts are normalized, log1p transformed and PCA is applied to select the first 50 PCs.
- *Leiden*: The genes with normalized variance computed in *Seurat v3* style larger than 1 are selected, normalized, log1p transformed, scaled and PCA is applied to select the first 15 PCs.

The considered hyperparameter values suggested by the authors, as well as those discussed in the corresponding paper can be found in table S1. For *Leiden*, we iteratively increase the resolution parameter until the desired number of clusters is obtained. We trained and evaluated methods using GPU when available; otherwise, we utilized CPU.

| Model | Hyperparameters |
|---|---|
| *BayesSpace* | model: [$normal, t$]; gamma: $[1, 2, 3]$ |
| *stLearn* | weights: [weights_matrix_all, weights_matrix_pd_gd, weights_matrix_pd_md, weights_matrix_gd_md, gene_expression_correlation, physical_distance, morphological_distance] |
| *GraphST* | alpha: $[1, 10, 20]$; beta: $[1, 10, 20]$; lambda1: $[1, 10, 20]$; lambda2: $[1, 10, 20]$; radius: $[0, 50]$ |
| *STAGATE* | k_cutoff: $[6, 9, 12, 15]$; hidden_dims: $[[512, 30], [256, 30], [512, 60], [256, 60]]$; pre_resolution: $[0.2, 0.4, 0.6]$ |
| *MUSE* | lambda_regul: $[1, 5, 10]$; lambda_super: $[1, 5, 10]$ |
| *SpaGCN* | s: $[1, 2, 3]$; histology: [$False, True$]; n_neighbors: $[5, 15, 30]$ |
| *AESTETIK* | morphology_weight: $[0, 1, 1.5, 2, 3]$; window_size: $[5, 7]$; refine_cluster: [$False, True$] |

**Table S1** Model hyperparameters.

| SampleID | Replicate Pair | Patient |
|---|---|---|
| 151507 | 1 | A |
| 151508 | 1 | A |
| 151509 | 2 | A |
| 151510 | 2 | A |
| 151669 | 1 | B |
| 151670 | 1 | B |
| 151671 | 2 | B |
| 151672 | 2 | B |
| 151673 | 1 | C |
| 151674 | 1 | C |
| 151675 | 2 | C |
| 151676 | 2 | C |

**Table S2** The LIBD Human DLPFC contains 3 patients (A, B, C) with two pairs of spatially adjacent replicates (1, 2) resulting in 12 tissue slices.

### 2 Data

#### 2.1 LIBD Human DLPFC

The LIBD Human DLPFC dataset [7] comprises 12 tissue slices obtained from the dorsolateral prefrontal cortex (DLPFC) brain region, sequenced using Visium from 10x Genomics™ (Table S2). Each spot is manually annotated based on brain cytoarchitecture and known marker genes.

#### 2.2 Human Breast Cancer

The Human Breast Cancer dataset [8] contains 6 tissue slices sequenced using Visium from 10x Genomics™ in 2 independent biological labs (Table S3). Due to incomplete and low-quality annotation, slice CID4290 is removed from the analysis. Each spot is manually annotated based on Loupe v.4.0.0 software (10x Genomics™).

| SampleID | Lab |
|---|---|
| CID4465 | A |
| CID44971 | A |
| CID4535 | A |
| CID4290 | A |
| 1142243F | B |
| 1160920F | B |

**Table S3** The Human Breast Cancer dataset contains 6 samples sequenced in 2 independent biological labs - A and B.

| Channel | Liver Normal | Liven Cancer |
|---|---|---|
| 1 | PanCK | PanCK |
| 2 | CK8/18 | CD68 |
| 3 | CD298/B2M | CD298/B2M |
| 4 | CD45 | CD45 |
| 5 | DAPI | DAPI |

**Table S4** Channel order and the corresponding segmentation marker in CosMx NanoString™ Human Liver dataset.

#### 2.3 Tumor Profiler

The metastatic melanoma dataset comprises 18 tissue slices sequenced using Visium from 10x Genomics™ from the Tumor Profiler study [9]. The tissue slices originate from 9 tissue regions (6.5 x 6.5 $mm^2$) from 7 donors, each characterized by one of the following immune subtypes: immune desert, immune excluded, or inflamed. It contains 1 replicate for each tissue region - resulting in 18 tissue slices. The H&E images have (or are scaled to) a resolution of 0.30µm/pixel with a spot radius of 160 pixels. The data is generated using 10x Genomics™ *Space Ranger v3.0.0*. The ground truth annotations were generated using histopathology software (HALO AI™ (Indica Labs, Corrales, NM, USA)), classifying the spots into one of the following categories: tumor, stroma, normal lymphoid, and blood/necrosis. Following that, the model predictions underwent manual review by a pathologist.

#### 2.4 CosMx NanoString™ Human Liver

The CosMx NanoString™ Human Liver dataset, derived from normal liver and hepatocellular carcinoma tissues, was produced using the CosMx Human Universal Cell Characterization RNA panel [10]. We used the provided raw Morphology2D Normalized TIFF images with 5 channels, corresponding to the expression level of segmentation markers (Table S4), to create an RGB image with 3 channels by applying PCA. The provided cell profiles are grouped into Field of Views (FOVs) of around 1,500 cells each along with their spatial coordinates, and cell type (304 FOVs normal, 383 FOVs cancer). The *Hep* cell groups were merged. Cells situated within a 55-pixel distance from the image boundaries are removed. For computational efficiency, we sample 100 FOVs per condition.

#### 2.5 Simulated data

We adapted the simulation approach suggested in [5] by introducing spatial structure in the experiment. Briefly, relying on simulated ground truth labels, we simulate transcriptomics and morphology modalities, allowing partial observation of true clusters within each modality individually. However, combining both modalities enables the identification of all clusters. Spatial coordinates are incorporated by sorting the ground truth in spatial space. We generated 3 datasets with 5 samples (2500 cells/sample) and 5, 10, and 15 clusters each. For the runtime analysis, we generated one tissue slice with 10 million spots and 5 clusters.

#### 2.6 Runtime analysis

We applied *AESTETIK* with morphology weight 1.5, window size 3 and a batch size 100 000. Due to the large number of spots, we used *K-Means* clustering from the *sklearn* package. The model was trained on NVIDIA™ V100 GPU with 32G RAM. We conducted runtime benchmarking for three key functions of *AESTETIK* v0.0.1: grid building (`prepare_input_for_model()`), training (`train()`), and inference (`compute_spot_representations()`).

### 3 Suplementary figures

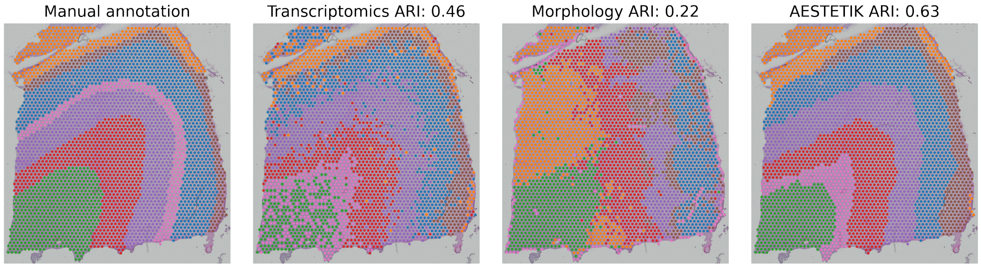


**Fig. S1 Comparison of cluster assignments for slice 151676 from the LIBD human DLPFC dataset - Transcriptomics only vs Morphology only vs *AESTETIK*.**

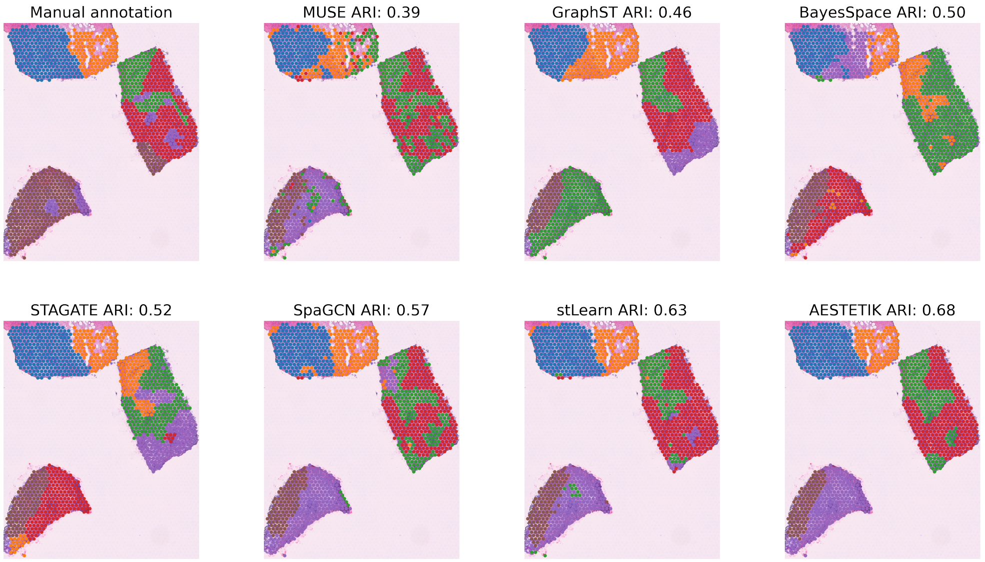


**Fig. S2 Comparison of cluster assignments for slice CID44971 from the Human Breast Cancer dataset.**

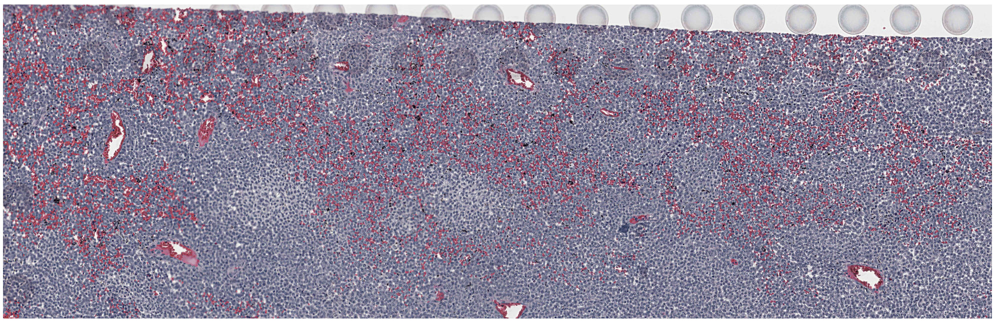

**Fig. S3 Zoom into the hemorrhage region highlighted in Fig. 3A, B for slice MACEGEJ-2-2.**

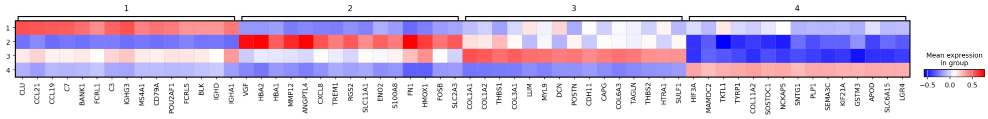


**Fig. S4 Cluster marker genes for slice MACEGEJ-2-2 from the Tumor Profiler dataset.** Cluster 1 corresponds to spots enriched with normal lymphoid cells; cluster 2 - blood and necrosis; cluster 3 - stroma; cluster 4 - tumor.

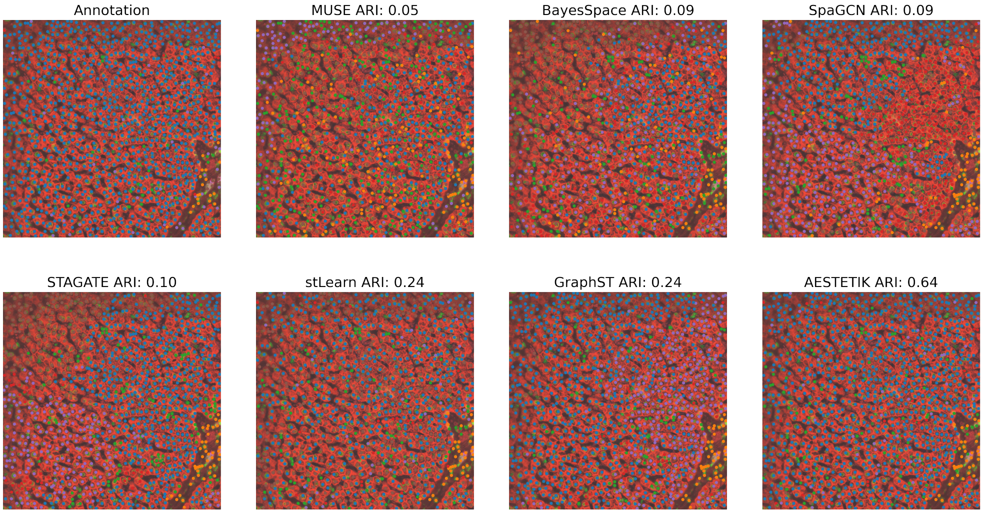


**Fig. S5 Comparison of cluster assignments for FOV 159 from the CosMx NanoString™ Liver Normal dataset.**

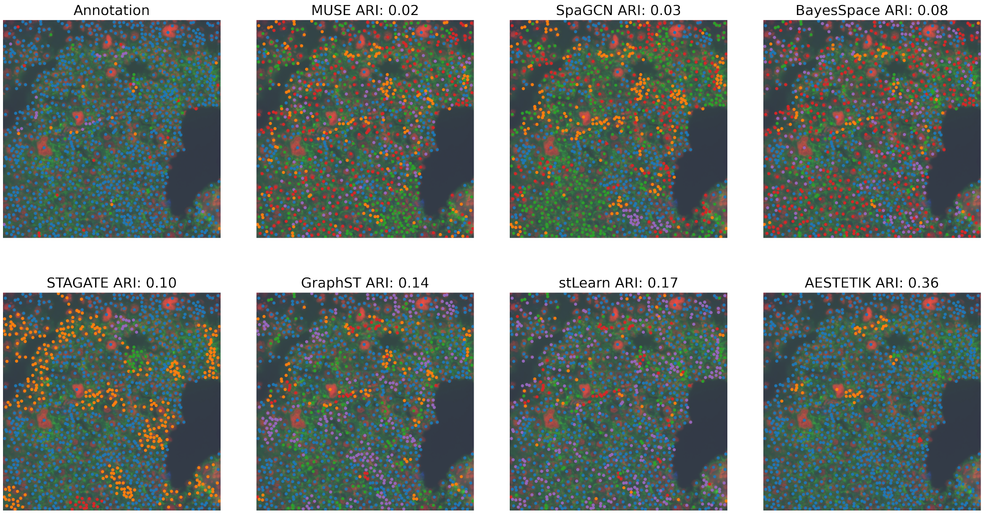


**Fig. S6 Comparison of cluster assignments for FOV 201 from the CosMx NanoString™ Liver Cancer dataset.**

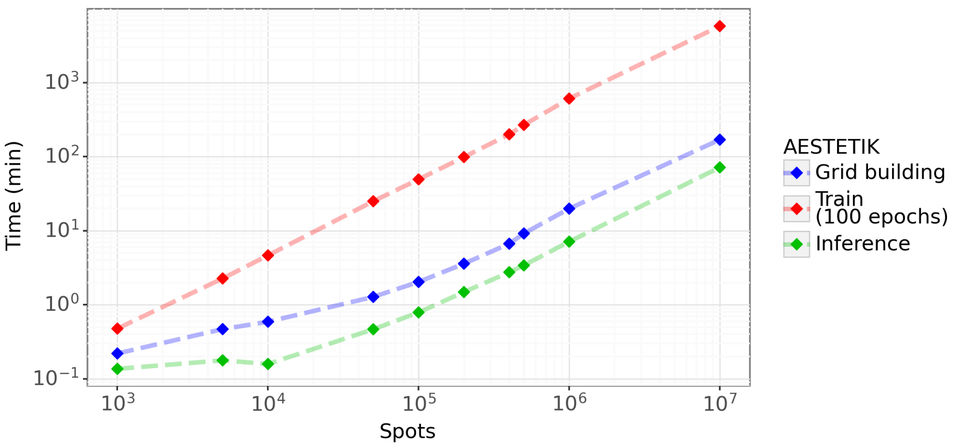


**Fig. S7** ***AESTETIK* scales to millions of spots.**
The x-axis represents the number of spots and the y-axis - the time in minutes.
